# Impact of changes in Achilles tendon thickening on cardiovascular events in patients with familial hypercholesterolemia

**DOI:** 10.1101/2023.12.29.23300597

**Authors:** Hayato Tada, Nobuko Kojima, Yasuaki Takeji, Atsushi Nohara, Masa-aki Kawashiri, Masayuki Takamura

## Abstract

**Background:** Achilles tendon thickening (ATT) can be ameliorated by lowering low-density lipoprotein (LDL) levels in patients with familial hypercholesterolemia (FH). The Japan Atherosclerosis Society (JAS) defines ATT as ≥8.0 mm in males and ≥7.5 mm in females. We aimed to determine the clinical impact of changes in ATT on the development of major adverse cardiovascular events (MACE).

**Methods:** Patients with clinically diagnosed heterozygous FH (HeFH) (N = 1273; 614 males, 659 females) with ATT data from X-ray were assessed. Patients were divided into four groups: patients without ATT from baseline until follow-up (group 1), patients without ATT at baseline but developed ATT at follow-up (group 2), patients with ATT at baseline but regressed at follow-up (group 3), and patients with ATT from baseline until follow-up (group 4). Cox proportional hazard models were used to assess the factors associated with MACE, including cardiovascular death and any coronary events.

**Results:** On follow-up (median: 10.9 years), 142 MACEs were observed, and the median ATT regressed from 7.8 to 7.6 mm. Changes in ATT were significantly associated with the occurrence of MACE in all groups, when compared to group 1 (hazard ratio [HR]: 2.73; 95% confidence interval [CI]: 1.33–4.13 [p < 0.001], HR: 2.18, 95% CI: 1.08–3.28, [p < 0.001], HR: 6.34, 95% CI: 3.10–9.58, [p < 0.001], in groups 2, 3, and 4, respectively).

**Conclusions:** Assessing ATT has diagnostic value and allows for risk stratification among patients with HeFH.

## 1. Introduction

Familial hypercholesterolemia (FH) is one of the most common Mendelian disorders, with an estimated prevalence of 1 in 300 among the general population [1–4]. This disorder has three major clinical manifestations, including high low-density lipoprotein (LDL) cholesterolemia, premature coronary artery disease (CAD), and tendon xanthomas [5], which are used as diagnostic criteria of FH worldwide [6–10]. In particular, tendon xanthomas, especially Achilles tendon thickening (ATT), are an important objective diagnostic criterion used by the Japan Atherosclerosis Society (JAS) because it can be assessed as a numerical value. In 2022, the JAS FH clinical criteria were revised; the thresholds of ATT on X-ray were changed from ≥9.0 mm in both sexes to ≥8.0 mm in males and ≥7.5 mm in females [9]. ATT can be ameliorated with LDL-lowering therapies, especially in younger patients [11], with some reaching normal levels but others that remained unchanged. In this study, we determined the clinical impact of changes in ATT on the development of major adverse cardiovascular events (MACE) among patients with heterozygous FH (HeFH).

## 2. Materials and Methods

### 2.1. Study population

Patients at Kanazawa University Hospital diagnosed with HeFH from 1990 to 2022 were retrospectively assessed. We used the most recent Japanese clinical criteria for FH (JAS FH criteria 2022). Out of 2213 HeFH patients, 1273 were included in the analysis, after excluding those with a lack of baseline clinical data (e.g., ATT, presence of CAD) and lost to follow-up.

### 2.2. Clinical data

We assessed baseline clinical variables, including age, sex, hypertension (systolic blood pressure ≥ 140 mmHg and/or diastolic blood pressure ≥ 90 mmHg, or receiving antihypertensive treatment), diabetes (according to the Japanese clinical diagnostic criteria of diabetes), smoking (defined as current smoking habit), total cholesterol, triglycerides, HDL cholesterol, LDL cholesterol (using the Friedewald formula if serum triglyceride levels were <400 mg/dL; otherwise, determined enzymatically), and the presence of CAD (severe stenosis [≥75%] of any coronary artery). ATTs were assessed by X-ray using the standard procedure at baseline and at follow-up [12]. Cholesterol-year scores were assessed as previously described [13]. We also assessed the occurrence of major adverse cardiac events (MACEs) during follow-up, defined as cardiovascular death or any coronary event, including coronary revascularization.

### 2.3. Changes in ATT status

Patients were divided into four groups based on changes in ATT status, defined as ≥8.0 mm in males and ≥7.5 mm in females. Group 1 included patients without ATT from baseline until follow-up. Group 2 included patients without ATT at baseline but developed ATT at follow-up. Group 3 included patients with ATT at baseline but regressed at follow-up. Group 4 included patients with ATT from baseline until follow-up.

### 2.4. Genetic analysis

The coding regions of FH genes, including LDL receptor, proprotein convertase subtilisin/kexin type 9 (*PCSK9*), apolipoprotein B (*APOB*), and LDL receptor adaptor protein 1 (*LDLRAP1*), were sequenced using iSeq 100 (San Diego, USA) as previously described [14]. The pathogenicity of the variants was determined using allele frequency information obtained from the ExAC Asian population data, *in silico* annotation tools, and the ClinVar database. Rare mutations in the Asian population were defined as those with an allele frequency <5%. Finally, the pathogenicity of the variants was classified according to the standard American College of Medical Genetics and Genomics criteria, as previously described [15].

### 2.5. Ethical considerations

This study was conducted in compliance with the Declaration of Helsinki, the Ethical Guidelines for Medical and Health Research Involving Human Subjects, and all other applicable laws and guidelines in Japan. The study protocol was approved by the Institutional Review Board of Kanazawa University Hospital (Kanazawa, Japan). Informed consent for genetic analyses was obtained from all patients.

### 2.6. Statistical analysis

Categorical variables were reported as percentages and compared using Fisher’s exact test or the chi-square test, as appropriate. Continuous variables with normal and nonnormal distributions, respectively, were reported as means ± SD and medians with interquartile ranges (IQRs). The mean values of continuous variables were compared using Student’s t-test for independent data variables, whereas median values were compared using the nonparametric Wilcoxon Mann–Whitney rank sum test. Chi-square or Fisher’s post hoc tests were used for categorical variables as indicated. Spearman’s correlation coefficients were calculated between age and Achilles tendon thickness and between cholesterol-year score and Achilles tendon thickness. Cox proportional hazards model was used to assess relationships among all variables, including age, sex, hypertension, diabetes, smoking status, baseline LDL cholesterol, prior CVD, FH mutations, and ATT. Cumulative Kaplan– Meier survival curves starting at baseline were constructed to compare the time to the first MACE incident. Statistical analysis was conducted using R statistics (https://www.r-project.org), with p < 0.05 considered statistically significant.

## 3. Results

### 3.1. Clinical characteristics at baseline based on the occurrence of MACE

Baseline clinical characteristics are presented in Table 1. The mean age was 50 years, with 614 (48.2%) males. The median LDL cholesterol at baseline was 241 mg/dL, which was reduced to 113 mg/dL at follow-up under the lipid-lowering therapies (**Supplemental Table 1**). The median Achilles tendon thickness at baseline was 7.8 mm, which was reduced to 7.6 mm at follow-up. There were significant differences in most variables, except for the prevalence of FH mutation, when patients were grouped according to the occurrence of MACE (Table 1). The cholesterol-year score was significantly associated with age (**Supplemental Figure 1)**.

**Table 1.** Baseline characteristics according to the occurrence of major adverse cardiovascular events.

|  | All<br>(N = 1,273) | MACE<br>(N = 142) | NO-MACE<br>(N = 1,131) | *P*-value |
| --- | --- | --- | --- | --- |
| Age (years) | 50 ± 16 | 61 ± 16 | 47 ± 16 | <0.001 |
| Male (%) | 614 (48.2%) | 80 (56.3%) | 534 (47.2%) | 0.0498 |
| Hypertension (%) | 368 (28.9%) | 102 (71.8%) | 266 (13.5%) | <0.001 |
| Diabetes (%) | 114 (9.0%) | 43 (30.3%) | 71 (6.3%) | <0.001 |
| Smoking (%) | 228 (17.9%) | 85 (59.9%) | 303 (26.8%) | <0.001 |
| Total cholesterol (mg/dL) | 317 [288–359] | 321 [289–377] | 317 [287–355] | <0.001 |
| Triglyceride (mg/dL) | 125 [83–173] | 136 [94–178] | 124 [82–170] | <0.001 |
| HDL cholesterol (mg/dL) | 47 [40–57] | 44 [35–52] | 48 [40–57] | < 0.001 |
| LDL cholesterol (at baseline, mg/dL) | 241 [208–286] | 253 [213–309] | 240 [207–283] | <0.001 |
| LDL cholesterol (at follow-up mg/dL) | 113 [94–135] | 110 [88–126] | 113 [95–135] | <0.001 |
| Family history of FH and/or premature CVD (%) | 920 (72.3%) | 124 (87.3%) | 896 (79.2%) | 0.03 |
| Achilles tendon thickness (at baseline, mm) | 7.8 [6.6–9.0] | 9.0 [8.0–13.3] | 7.6 [6.6–9.0] | <0.001 |
| Achilles tendon thickness (at follow-up, mm) | 7.6 [6.4–8.9] | 9.0 [8.0–13.0] | 7.5 [6.4–8.9] | <0.001 |
| FH mutation (%) | 966 (75.9%) | 112 (78.9%) | 854 (75.5%) | 0.44 |
| History of prior CVD (%) | 393 (30.9%) | 115 (81.0%) | 278 (24.6%) | <0.001 |

HDL, high-density lipoprotein; LDL, low-density lipoprotein; CVD, cardiovascular disease

### 3.2. Associations between ATT and clinical variables

Achilles tendon thickness was significantly larger in patients with MACE versus those without (median: 9.0 vs. 7.6 mm, **Figure 1A, Supplemental Figure 2**), as well as in patients with FH mutation versus those without (8.0 vs. 6.6 mm, **Supplemental Figure 3**). In addition, Achilles tendon thickness increased with aging among patients with HeFH in both males (p < 0.001, Spearman’s r = 0.22) and females (p <0.001, Spearman’s r=0.31). Similarly, Achilles tendon thickness among patients with HeFH was correlated with cholesterol-year score regardless of the occurrence of MACE (p < 0.001; Spearman’s r = 0.33 in patients with MACE, and p < 0.001; Spearman’s r = 0.35 in patients without MACE, **Figure 2**)

**Figure 1:**
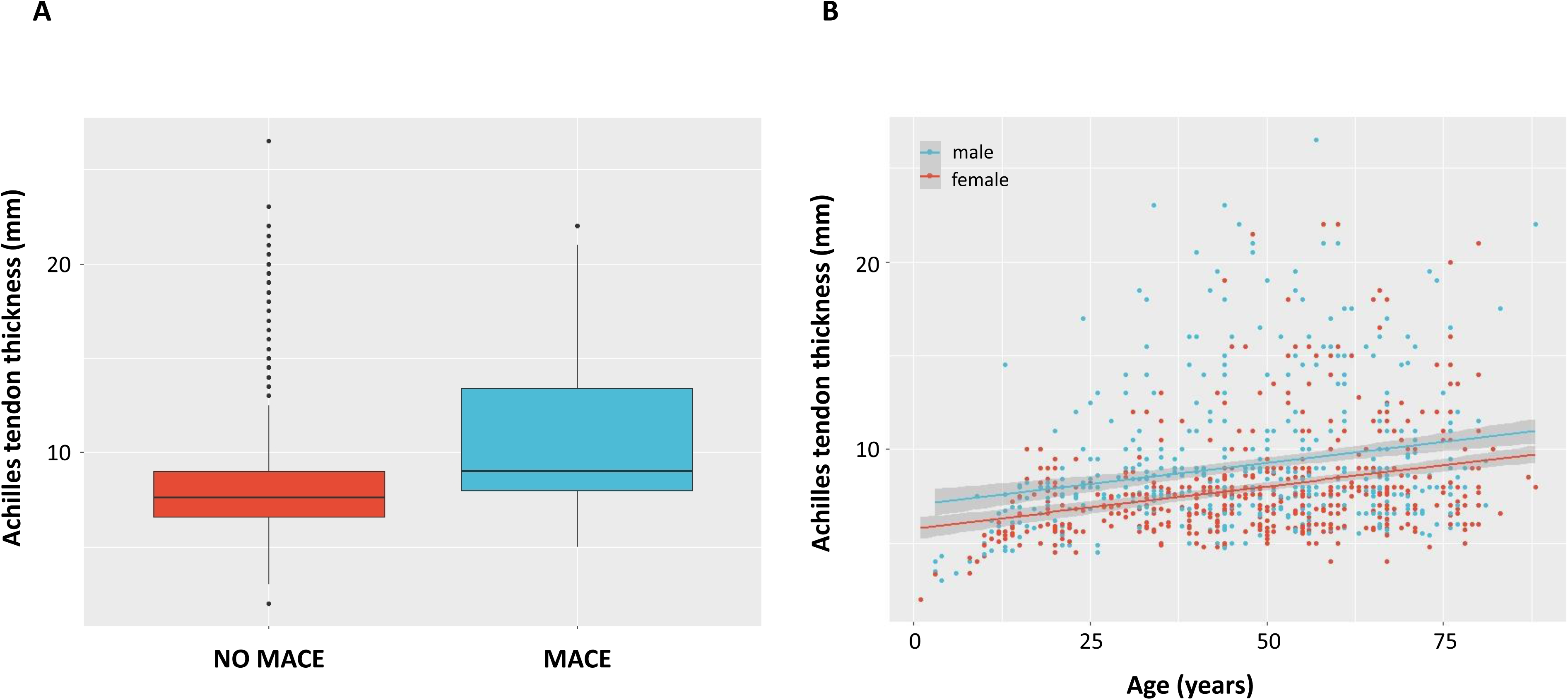
Association between Achilles tendon thickness and major adverse cardiovascular events (MACE) and age. A. Box plot of Achilles tendon thickness according to the presence of MACE Red: patients without MACE. Green: patients with MACE. B. Correlation between Achilles tendon thickness and age Red indicates female. Green indicates male.

**Figure 2:**
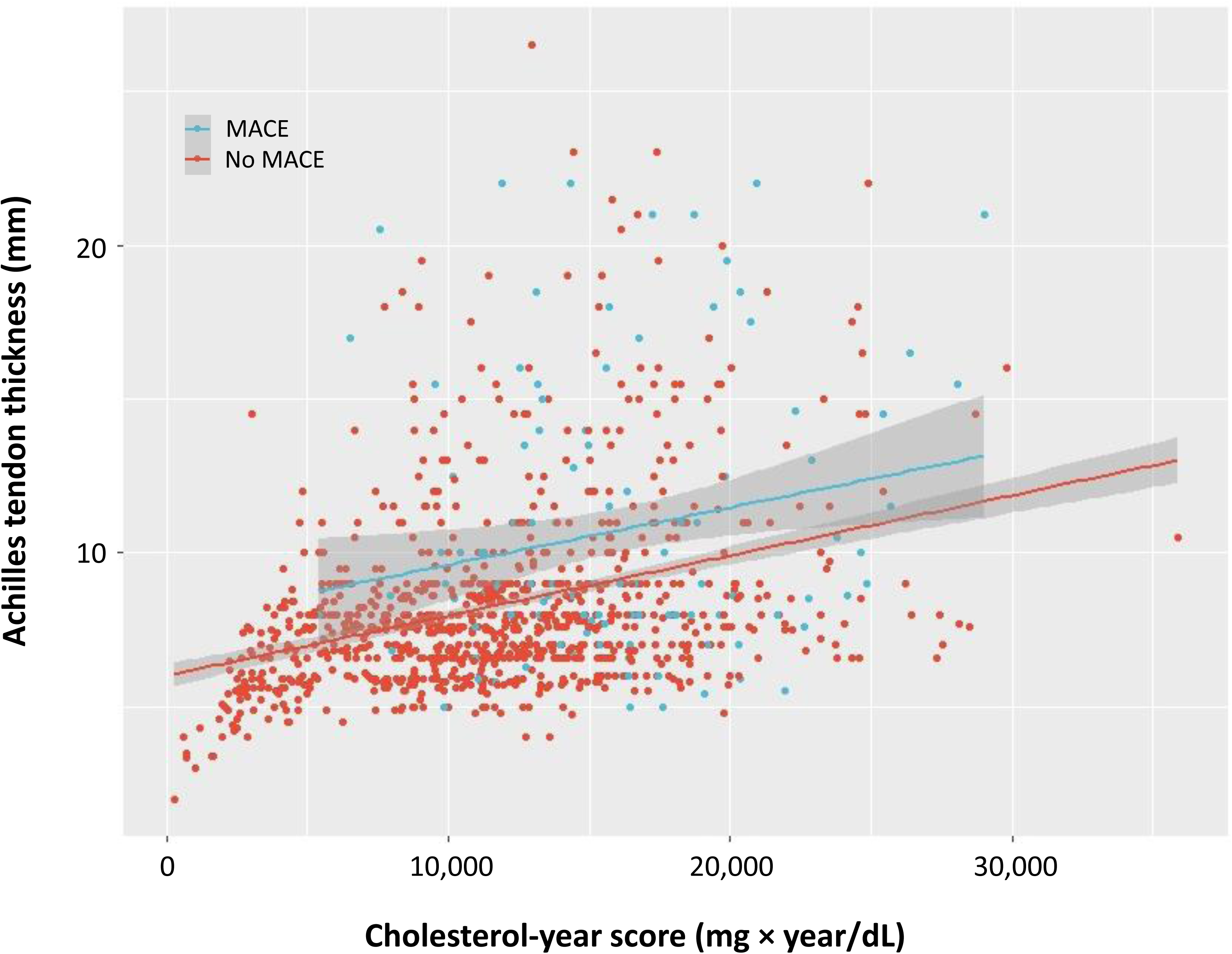
Correlation between Achilles tendon thickness and the cholesterol-year score. Red: patients without major adverse cardiovascular events (MACE). Green: patients with MACE.

### 3.3. Baseline and changes in Achilles tendon thickness according to age

There was a significant trend between age and Achilles tendon thickness (**Figure 3A**). The median (IQR) Achilles tendon thickness of each age group was 4.0 mm (3.4–4.3) in 0–9 years, 6.6 mm (5.5–7.4) in 10–19 years, 7.4 mm (6.0–8.2) in 20–29 years, 8.0 mm (6.8–9.0) in 30–39 years, 7.8 mm (6.6–9.0) in 40–49 years, 7.7 mm (6.6–9.5) in 50–59 years, 8.0 mm (6.8–11.0) in 60–69 years, 8.4 mm (7.0–11.0) in 70–79 years, and 8.5 mm (range,7.6–11.0) in 80–89 years. Larger regressions in Achilles tendon thickness were observed in younger generations (**Figure 3B**). The median (IQR) changes in Achilles tendon thickness of each age group were +0.5 mm (+0.2–+1.0) in 0–9 years, −0.2 mm (−0.6–+0.1) in 10–19 years, −0.2 mm (−1.0–+0.1) in 20–29 years, −0.2 mm (−1.0–0.0) in 30–39 years, −0.1 mm (−0.8–+0.1) in 40–49 years, 0.0 mm (−0.5–+0.2) in 50–59 years, 0.0 mm (0.0–+0.4) in 60–69 years, 0.0 mm (0.0–+0.4) in 70–79 years, 0.0 mm (0.0–+0.3) in 80–89 years.

**Figure 3:**
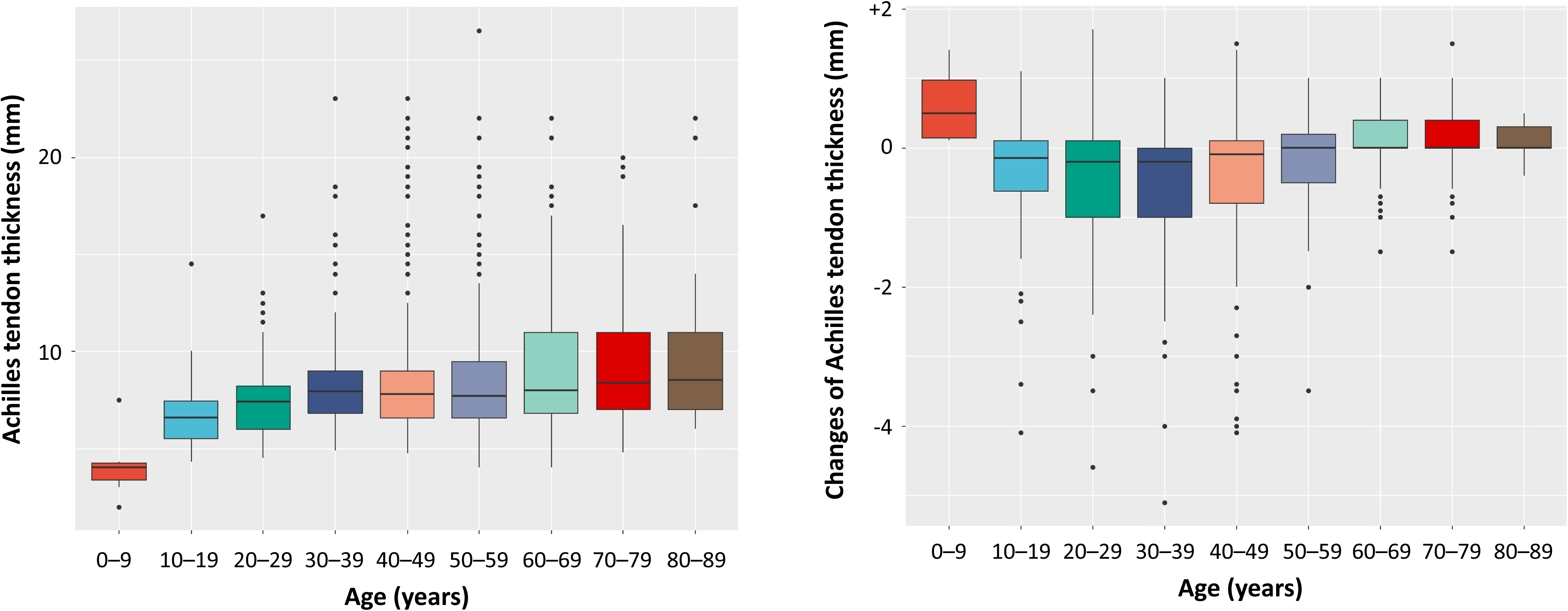
Baseline and changes in Achilles tendon thickness according to age. A. Boxplots of Achilles tendon thickness according to age B. Boxplots of changes in Achilles tendon thickness according to age

### 3.4. Clinical characteristics at baseline according to changes in ATT

Some patients without ATT at baseline remained without ATT (group 1, N = 158), while others developed ATT during follow-up (group 2, N = 91). Moreover, there were also some patients with ATT at baseline whose ATT regressed at follow-up (group 3, N = 111) and others whose ATT remained unchanged during follow-up (group 4, N = 913). Detailed clinical characteristics according to this classification are described in **Table 2**.

**Table 2.** Baseline characteristics according to changes in Achilles tendon thickening status.

|  | All<br>(N = 1,273) | group 1<br>(N = 158) | group 2<br>(N = 91) | group 3<br>(N = 111) | group 4<br>(N = 913) |
| --- | --- | --- | --- | --- | --- |
| Age (years) | 50 ± 16 | 46 ± 18 | 53 ± 16 | 42 ± 18 | 50 ± 16 |
| Male (%) | 614 (48.2%) | 66 (41.8%) | 54 (59.3%) | 54 (48.6%) | 440 (48.2%) |
| Hypertension (%) | 368 (28.9%) | 31 (19.6%) | 37 (40.6%) | 34 (30.6%) | 266 (29.1%) |
| Diabetes (%) | 114 (9.0%) | 10 (6.3%) | 12 (13.2%) | 6 (5.4%) | 86 (9.4%) |
| Smoking (%) | 228 (17.9%) | 15 (9.5%) | 31 (34.1%) | 11 (10.0%) | 171 (18.7%) |
| Total cholesterol (mg/dL) | 317 [288–359] | 311 [266–351] | 321 [276–363] | 325 [291–391] | 317 [287–358] |
| Triglyceride (mg/dL) | 125 [83–173] | 103 [74–156] | 134 [91–196] | 125 [81–165] | 124 [85–174] |
| HDL cholesterol (mg/dL) | 47 [40–57] | 50 [43–60] | 44 [33–51] | 48 [41–58] | 47 [40–57] |
| LDL cholesterol (at baseline, mg/dL) | 241 [208–286] | 238 [203–282] | 244 [213–294] | 255 [218–312] | 241 [210–286] |
| LDL cholesterol (at follow-up mg/dL) | 113 [94–135] | 110 [88–126] | 134 [98–152] | 104 [72–104] | 113 [95–135] |
| Family history of FH and/or premature CVD (%) | 920 (72.3%) | 104 (65.8%) | 67 (73.6%) | 68 (61.3%) | 681 (74.6%) |
| Achilles tendon thickening (at baseline, mm) | 7.8 [6.6–9.0] | 6.9 [6.2–7.4] | 7.1 [6.8–7.5] | 8.6 [7.6–10.0] | 8.8 [8.1–13.6] |
| Achilles tendon thickening (at follow-up, mm) | 7.6 [6.4–8.9] | 6.8 [6.1–7.3] | 8.4 [7.9–9.4] | 7.2 [7.4–10.0] | 8.8 [8.0–13.5] |
| FH mutation (%) | 966 (75.9%) | 72 (45.6%) | 77 (84.6%) | 104 (93.7%) | 713 (78.1%) |
| History of prior CVD (%) | 393 (30.9%) | 15 (9.5%) | 26 (28.6%) | 36 (32.4%) | 316 (34.6%) |

HDL, high-density lipoprotein; LDL, low-density lipoprotein; CVD, cardiovascular disease; FH, familial
hypercholesterolemia

### 3.5. Factors associated with MACE

In total, 142 MACEs were observed during the median follow-up period of 10.9 years (IQR, 7.8–15.4 years). The details of MACEs are described in **Supplemental Table 2**. The Cox proportional hazards model revealed that the following factors were significantly associated with MACEs: age (hazard ratio [HR] = 1.07; 95% confidence interval [CI] = 1.05–1.10; *p* < 0.001), male sex (HR = 1.55; 95% CI = 1.08–2.10; *p* < 0.001), hypertension (HR = 3.14; 95% CI = 2.00–4.28; *p* < 0.001), diabetes (HR = 2.54; 95% CI = 1.50–3.58; *p* < 0.001), smoking (HR = 2.22; 95% CI = 1.50–2.94; *p* < 0.001), LDL cholesterol (HR = 1.01; 95% CI = 1.00–1.02; *p* < 0.001, per 10 mg/dL), prior CVD (HR = 3.11; 95% CI = 1.88–4.54; *p* < 0.001), and the presence of pathogenic variants (HR = 1.66; 95% CI = 1.11–2.21; *p* < 0.001) (**Table 3**). Under these circumstances, changes in ATT were significantly associated with the occurrence of MACE, in group 2 (HR: 2.73; 95% CI: 1.33–4.13; p < 0.001), group 3 (HR: 2.18; 95% CI: 1.08–3.28; p < 0.001), and group 4 (HR: 6.34; 95% CI: 3.10–9.58; p < 0.001), when group 1 was considered as reference.

**Table 3.** Factors associated with major adverse cardiovascular events.

| Variable | HR | 95% CI | *P*-value |
| --- | --- | --- | --- |
| Age (per year) | 1.07 | 1.05–1.10 | <0.001 |
| Male (yes vs. no) | 1.55 | 1.08–2.10 | <0.001 |
| Hypertension (yes vs. no) | 3.14 | 2.00–4.28 | <0.001 |
| Diabetes (yes vs. no) | 2.54 | 1.50–3.58 | <0.001 |
| Smoking (yes vs. no) | 2.22 | 1.50–2.94 | <0.001 |
| LDL cholesterol at baseline (per 10 mg/dl) | 1.01 | 1.00–1.02 | <0.001 |
| Prior CVD (yes vs. no) | 3.11 | 1.88–4.54 | <0.001 |
| FH mutation (yes vs. no) | 1.66 | 1.11–2.21 | <0.001 |
| ATT at baseline ($\geq 8.0$ mm in males, $\geq 7.5$ mm in females) | 2.44 | 1.36–3.46 | <0.001 |
| Group 2 (Group 1 as reference) | 2.73 | 1.33–4.13 | <0.001 |
| Group 3 (Group 1 as reference) | 2.18 | 1.08–3.28 | <0.001 |
| Group 4 (Group 1 as reference) | 6.34 | 3.10–9.58 | <0.001 |

LDL, low-density lipoprotein; CVD, cardiovascular disease; FH, familial hypercholesterolemia; ATT, Achilles tendon
thickening

### 3.6. The occurrence of MACE according to changes in ATT status

Group 4 had the worst prognosis, followed by groups 2 and 3. The prognosis of group 1 was quite good (**Figure 4**).

**Figure 4.**
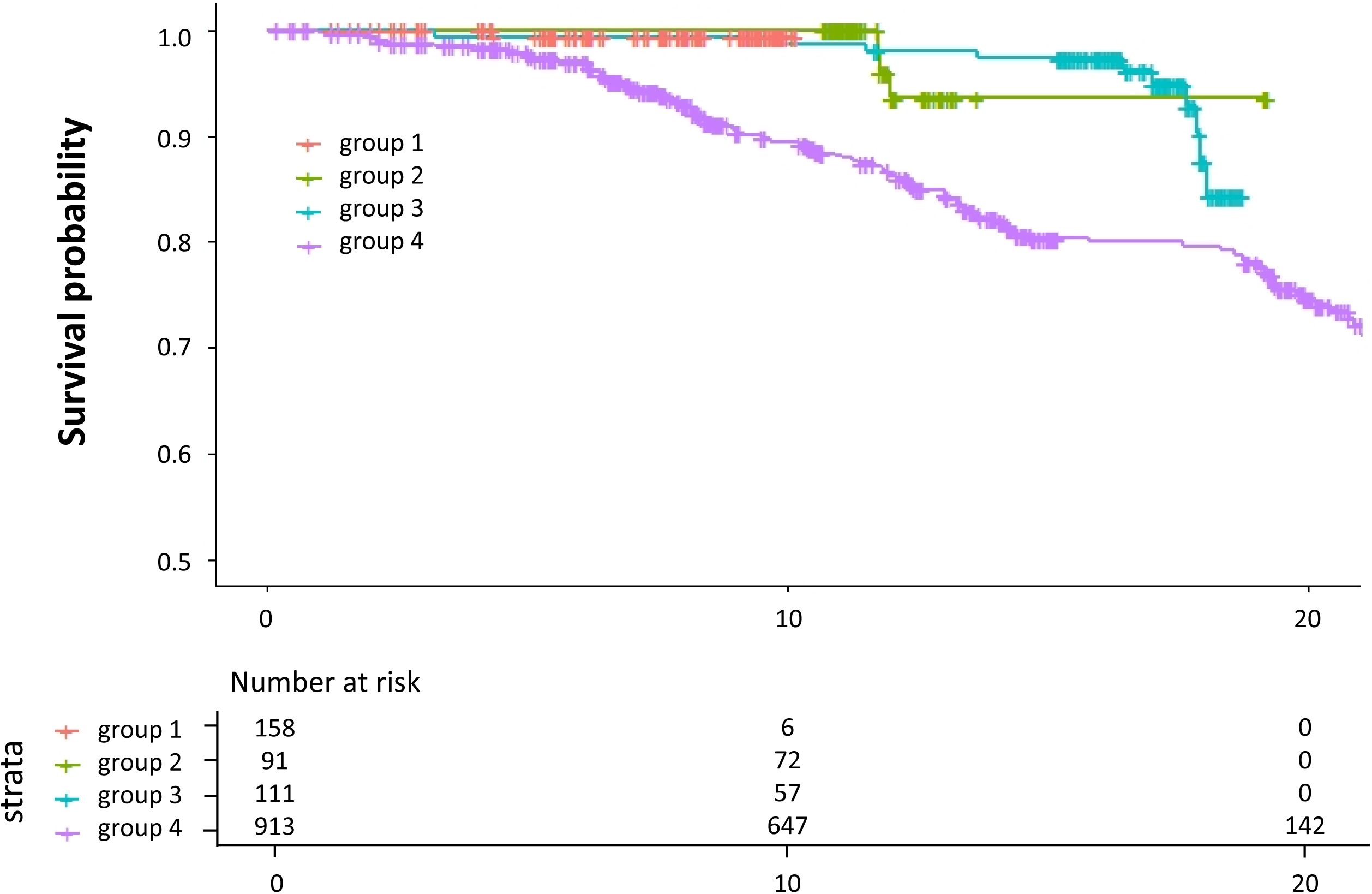
Kaplan–Meier survival curve. The x-axis represents the follow-up period (years). The y-axis represents the proportion of patients without major adverse cardiovascular events (MACE). Red: group 1. Green: group 2. Blue: group 3. Purple: group 4.

## 4. Discussion

In this study, we determined the clinical impact of changes in ATT on the development of MACE. Our key findings were as follows: 1) Achilles tendon thickness in HeFH is associated with aging and cholesterol-year score, 2) ATT can regress in some patients, associated with LDL-lowering therapy, and 3) changes in ATT status can be used as a risk marker for cardiovascular events among patients with HeFH.

Tendon xanthomas, including ATT, is a specific finding of FH, and thus, it has been used as an important clinical diagnostic criteria of FH worldwide [6–10]. In fact, ATT can be objectively assessed via X-ray or ultrasound. Therefore, it is important to determine the threshold of ATT. In this study, we have decided to use the thresholds of ATT on X-ray as ≥8.0 mm in males and ≥7.5 mm in females, based on a multicenter study in Japan [12]. Using these cutoffs, patients with HeFH can be diagnosed more accurately with higher sensitivity [16]. We hope that the objective assessment of ATT will be used as a clinical diagnostic criteria in other countries. On the other hand, we noticed that ATT can regress in some patients associated with LDL-lowering therapies, especially younger patients [17]. However, it is unclear whether the assessments of ATT can also be used for risk stratification for cardiovascular events. In this study, we have clearly demonstrated that ATT assessments are useful even after follow-up. This is important because ATT can be assessed noninvasively. Although the mechanisms behind regression of ATT are still unclear, there are several implications regarding this topic. First, we observed more chances of ATT regression in younger patients. This is compatible with the notion that younger plaques with less fibrosis can regress under LDL-lowering therapies [18]. Second, we observed an association between FH mutation and ATT in this study. Rare genetic variants (e.g., FH mutations) appear to have a huge impact on the development of ATT.

This study has several limitations. First, we used retrospective data from a single center. Thus, the observations from this study may not be applicable to other studies. Second, functional analyses to validate the pathogenicity of FH mutations were not conducted. Third, the timing of assessments of ATT, especially at follow-up, was not systematically determined.

In conclusion, assessments of ATT carry diagnostic value and can be used for risk stratification information among patients with HeFH.

## Data Availability

All data produced in the present study are available upon reasonable request to the authors

## Acknowledgements

We are thankful to Ms. Kazuko Honda and Mr. Sachio Yamamoto for their outstanding technical assistance.

## Author contributions

The authors confirm their contribution to the paper as follows: Study conception: HT and MK; data collection: HT, NK, YT, AN, MK, and MT. Analysis and interpretation of results: HT and MK; draft manuscript preparation: HT, NK, YT, AN, MK, and MT. All authors have reviewed the results and approved the final version of the manuscript.

